# Performance characteristics of five antigen-detecting rapid diagnostic test (Ag-RDT) for SARS-CoV-2 asymptomatic infection: a head-to-head benchmark comparison

**DOI:** 10.1101/2021.02.11.21251553

**Authors:** Bàrbara Baro, Pau Rodo, Dan Ouchi, Antoni E. Bordoy, Emilio N. Saya Amaro, Sergi V. Salsench, Sònia Molinos, Andrea Alemany, Maria Ubals, Marc Corbacho-Monné, Pere Millat-Martinez, Michael Marks, Bonaventura Clotet, Nuria Prat, Jordi Ara, Martí Vall-Mayans, Camila G-Beiras, Quique Bassat, Ignacio Blanco, Oriol Mitjà

## Abstract

**Background:** Mass testing for early identification and isolation of infectious COVID-19 individuals, irrespective of concurrent symptoms, is an efficacious strategy to reduce disease transmission. Antigen-detecting rapid diagnostic tests (Ag-RDT) appear as a potentially suitable tool for mass testing on account of their ease-of-use, fast turnaround time, and low cost. However, benchmark comparisons are scarce, particularly in the context of unexposed asymptomatic individuals.

**Methods:** We used nasopharyngeal specimens from unexposed asymptomatic individuals to assess five Ag-RDTs: PanBio^™^ COVID-19 Ag Rapid test (Abbott), CLINITEST® Rapid COVID-19 Antigen Test (Siemens), SARS-CoV-2 Rapid Antigen Test (Roche Diagnostics), SARS-CoV-2 Antigen Rapid Test Kit (Lepu Medical), and COVID-19 Coronavirus Rapid Antigen Test Cassette (Surescreen). Samples were collected between December 2020-January 2021 during the third wave of the epidemic in Spain.

**Findings:** The analysis included 101 specimens with confirmed positive PCR results and 185 with negative PCR. For the overall sample, the performance parameters of Ag-RDTs were as follows: Abbott assay, sensitivity 38·6% (95% CI 29·1–48·8) and specificity 99·5% (97–100%); Siemens, sensitivity 51·5% (41·3–61·6) and specificity 98·4% (95·3–99·6); Roche, sensitivity 43·6% (33·7–53·8) and specificity 96·2% (92·4–98·5); Lepu, sensitivity 45·5% (35·6–55·8) and specificity 89·2% (83·8–93·3%); Surescreen, sensitivity 28·8% (20·2–38·6) and specificity 97·8% (94·5–99·4%). For specimens with cycle threshold (Ct) <30 in RT-qPCR, all Ag-RDT achieved a sensitivity of at least 70%, with Siemens, Roche, and Lepu assays showing sensitivities higher than 80%. In models according to population prevalence, all Ag-RDTs will have a NPV >99% and a PPV<50% at 1% prevalence.

**Interpretation:** Two commercial, widely available assays can be used for SARS-CoV-2 antigen testing to achieve sensitivity in specimens with a Ct<30 and specificity of at least 80% and 96%, respectively. Estimated negative and positive predictive values suggests the suitability of Ag-RDTs for mass screenings of SARS-CoV-2 infection in the general population.

**Funding:** Blueberry diagnostics, Fundació Institut d’Investigació en Ciències de la Salut Germans Trias i Pujol, and #YoMeCorono.org crowdfunding campaign.

**Research in context:** *Evidence before this study:* In December 2020, we searched on PubMed for articles containing the terms “antigen”, “test” (or Ag-RDT), and “SARS-CoV-2” or “COVID-19” either in the title or the abstract. Our search yielded 79 entries corresponding to articles written in English. Of them, 33 were articles presenting the diagnostic performance of qualitative lateral-flow antigen-detecting rapid diagnostic tests (Ag-RDT). Four of these articles reported the results of head-to-head comparisons of various Ag-RDTs; in all cases, the number of tests was lower than the recommended for retrospective assessments of diagnostic performance (i.e., minimum of 100 PCR positive and 100 PCR negative). Furthermore, all head-to-head comparisons found in the literature included specimens obtained among individuals with varying disease status (none of which asymptomatic), thus limiting the adequacy of the estimates for an asymptomatic screening strategy.

*Added value of this study:* We compared for the first time head-to-head five Ag-RDT using a powered set of fresh respiratory specimens PCR-confirmed positive or negative, collected from unexposed asymptomatic individuals during screening campaigns for early detection of SARS-CoV-2 infection. The sample size was large enough to draw robust conclusions. Our analysis identified four Ag-RDTs (i.e., assays marketed by Abbott, Siemens, Roche, and Surescreen) with specificity higher than 96%. Despite the low sensitivity for the overall sample (range 29% to 51%), the corresponding values for the subset of samples with Ct <30 were higher than 80% for Siemens, Roche, and Lepu assays. The estimated NPV for a screening performed in an area with 1% prevalence would be >99% for all tests, while the PPV would be <50%.

*Implications of all the available evidence:* Current data on the diagnostic performance of Ag-RDTs is heterogeneous and precludes benchmark assessments. Furthermore, the screening of asymptomatic populations is currently not considered among the intended uses of Ag-RDT, mostly because of lack of evidence on test performance in samples from unexposed asymptomatic individuals. Our findings add to the current evidence in two ways: first, we provide benchmarking data on Ag-RDTs, assessed head-to-head in a single set of respiratory specimens; second, we provide data on the diagnostic performance of Ag-RDTs in unexposed asymptomatic individuals. Our findings support the idea that Ag-RDTs can be used for mass screening in low prevalence settings and accurately rule out a highly infectious case in such setting.

## Introduction

Mass testing for early identification and isolation of individuals infected with the severe acute respiratory syndrome coronavirus 2 (SARS-CoV-2), irrespective of symptoms, is potentially an efficacious strategy to reduce disease transmission.^1^ Recent advances on the validation of Antigen-detecting Rapid Diagnostic Tests (Ag-RDTs) show promise to replace central laboratory techniques for epidemiological control of the SARS-CoV-2 through mass testing.

Reverse transcription-polymerase chain reaction (RT-qPCR) is the current gold standard for identifying the presence of the SARS-CoV-2 in respiratory specimens.^2^ More recently, transcription-mediated amplification (TMA) of the SARS-CoV-2 genome has been added to the repertoire of nucleic acid amplification tests (NAAT) for SARS-CoV-2 detection.^3^ Despite their high sensitivity, NAATs are associated with drawbacks that limit their use for community-based testing strategies, including the need for laboratory-processing, high cost, and long turnaround from sampling to results release.

Ag-RDTs, commonly used in diagnosing other infectious diseases, have emerged as an alternative tool that meets the requirements for frequent testing at the point-of-care: rapid turnaround time, low cost, and ease-of-use.^4^ Overall, Ag-RDTs have lower sensitivity than NAATs; however, clinical validation studies have consistently reported increasing sensitivities in specimens with higher viral loads. These findings, along with the growing body of evidence on the lack of infectivity of cases with low viral load,^5–8^ and the potential long tail of positivity when using highly sensitive methods such as PCR, suggest that frequent testing with Ag-RDTs―even those with low sensitivity―may be more effective than less frequent testing with RT-qPCR or TMA for mass screening campaigns to improve SARS-CoV-2 control.^8,9^

The performance parameters of Ag-RDTs are mostly based on testing respiratory specimens from clinically suspected cases^10–13^ and contacts after exposure to a positive case.^14–17^ However, the sensitivity bias associated with the viral load leads to high heterogeneity in the reported performance parameters, which strongly depend on the disease status and potential exposure (e.g., symptomatic vs. asymptomatic, contact vs. unexposed) of tested individuals. This heterogeneity precludes comparative analyses between tests assessed in different studies and challenges benchmarking of Ag-RDTs. Furthermore, head-to-head comparisons are scarce, particularly in samples from asymptomatic individuals, the target population of community-based screening strategies.^18,19^ In this study, we used fresh nasopharyngeal samples collected in routine mass screening campaigns of unexposed asymptomatic individuals to perform a head-to-head comparison of five Ag-RDTs.

## Methods

### Study design

As part of the surveillance program for pandemic control in Catalonia (North-East Spain), the local government launched NAAT-based systematic screenings in areas at high risk of an outbreak. The University Hospital Germans Trias i Pujol processed nasopharyngeal specimens collected in a healthcare area in North-East Spain (i.e., Metropolità Nord) with a catchment population of ∼1,400,000 people. These samples enabled us to assess the Ag-RDTs in line with The Foundation for Innovative New Diagnostics (FIND) target product profile for lateral flow assays that directly detect antigens of SARS-CoV-2 antigen assays,^20^ which recommends at least 100 known negative samples and 100 known positive samples with a documented RT-PCR result. In this study, we used samples collected between December 2020 and January 2021 (i.e., during the third wave of the epidemic in Spain) with RT-qPCR results available (i.e., data on cycle threshold [Ct]) to perform a head-to-head assessment of five Ag-RDTs. Samples with invalid results in any of the assessed Ag-RDTs were excluded from the analysis.

All samples used in this analysis had been collected in the setting of a public health surveillance program, and data were handled according to the General Data Protection Regulation 2016/679 on data protection and privacy for all individuals within the European Union and the local regulatory framework regarding data protection. The study protocol was approved by the ethics committee of Hospital Germans Trias i Pujol (Badalona, Spain).

### Procedures

Samples consisted of nasopharyngeal swabs collected by health care workers during mass testing of unexposed asymptomatic individuals living in areas at high risk of an outbreak. Swab specimens were placed into sterile tubes containing viral transport media (DeltaSwab Virus, Deltalab; or UTM Universal Transport Medium, Copan). The reference test (i.e., RT-qPCR) was performed on fresh samples stored at 2 – 8 °C for up to 24 hours; samples were then stored up to 12h at 2-8 °C until their use for the five Ag-RDTs.

RNA for RT-qPCR tests were extracted from fresh samples using the viral RNA/Pathogen Nucleic Acid Isolation kit for the Microlab Starlet or Nimbus platforms (Hamilton, USA), according to the manufacturer’s instructions. PCR amplification was conducted according to the recommendations of the 2019-nCoV RT-qPCR Diagnostic Panel of the Centers for Disease Control and Prevention (CDC) (REF), using the Allplex™ 2019-nCoV assay (Seegene, South Korea) on the CFX96 (Bio-Rad, USA) in line with manufacturer’s instruction. Briefly, a 25 μL PCR reaction mix was prepared that contained 8 μL of each sample’s nucleic acids, 2019-nCoV positive and negative controls, 5 μL of 2019-nCoV MOM (primer and probe mix) and 2 μL of real-time one-step Enzyme. Thermal cycling was performed at the following conditions: 20 min at 50 °C for reverse transcription, followed by 15 min at 95°C, and then 45 cycles of 15 sec at 94°C and 30 sec at 58°C. An RT-qPCR was considered positive according to the manufacturer’s instructions.^21^

Index tests included the following Ag-RDTs: PanBio^™^ COVID-19 Ag Rapid test (Abbott), CLINITEST® Rapid COVID-19 Antigen Test (Siemens), SARS-CoV-2 Rapid Antigen Test (Roche Diagnostics), SARS-CoV-2 Antigen Rapid Test Kit (Lepu Medical), and COVID-19 Coronavirus Rapid Antigen Test Cassette (Surescreen). Supplementary Table 1 provides further details regarding the specifications of each test. All Ag-RDT determinations were performed in parallel by two blinded technicians, who used approximately 100 μL of 1:2 mix of each kit buffer and the sample previously homogenized. Samples were applied directly to the test cassette and incubated for 15 minutes at room temperature before reading results at the naked eye, according to the manufacturer instructions (i.e., the presence of any test line (T), no matter how faint, indicates a positive result).

### Outcomes and statistical analysis

We calculated that a sample size of at least 73 positive specimens and 165 negative specimens would give 80% power to estimate overall sensitivity and specificity of Ag-RDT assays in our study. We based our calculation on the expected sensitivity and specificity in asymptomatic population of 65% and 96%,^16,22^ respectively, fixed precision of the point estimate of 2.5%, and confidence level of 95%. The calculation was in line with FIND recommendations for assessing Ag-RDTs that retrospective assessments should include a minimum of 100 samples per RT-PCR result.^20^

The primary analysis of the head-to-head comparison was the sensitivity and specificity of each Ag-RDT. Sensitivity and specificity were calculated as defined by Altman et al.,^23^ and reported as a percentage and the exact binomial 95% confidence interval (CI). Sensitivity was also analysed in a subset of samples with Ct<30, considered at high risk of transmission.

Secondary analyses were done assessing discordance between results obtained in each Ag-RDTs. Positive and negative-predictive values for each Ag-RDT at population prevalence between 1% and 15% for SARS-CoV-2 infection were modelled^24^ and plotted with the exact binomial 95% CI.^25^ All analyses and plots were performed using R version 3·6.^26^

### Role of the funding source

The funders of the study had no role in the study conception, design, conduct, data analysis, or writing of the report. All authors had full access to all the data in the study and had final responsibility for the decision to submit for publication.

## Results

Our sample collection included 316 fresh nasopharyngeal swabs from unexposed asymptomatic individuals who had a RT-qPCR result available. Of these, 30 were excluded because of lack of documented Ct value (n=25), incomplete results due to limited sample volume (n=1), or invalid results in any of the Ag-RDTs (n=4, all of them in the Lepu assay), resulting in a study set of 286 samples: 101 (35·3%) with positive RT-qPCR result and 185 (64·7%) with negative RT-qPCR result (Figure 1).

**Figure 1.**
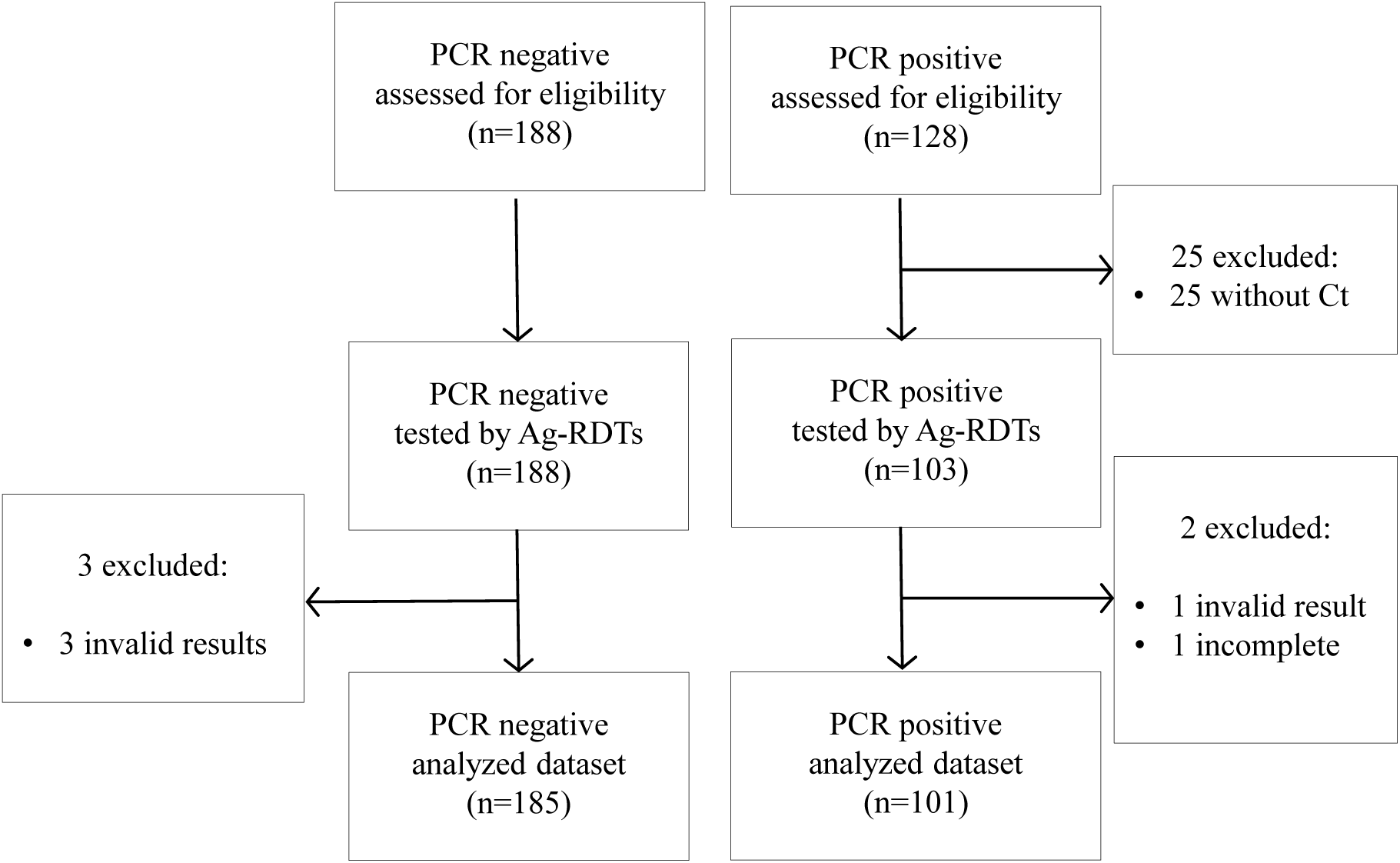
Flow-chart of sample inclusion. All samples were nasopharyngeal swabs collected from unexposed asymptomatic individuals during screening campaigns.

The Ct value of samples with positive RT-qPCR result was <30 in 30 (29·7%) samples, 30-to-35 in 46 (45·5%), and >35 in 25 (24·8%). The overall sensitivity and specificity of the analysed Ag-RDTs ranged from 28·7% to 51·5% and 89·2% to 99·5%, respectively (Table 1). When considering only RT-qPCR positive samples with Ct <30 (i.e., indicates a high concentration of viral genetic material which is typically associated with a higher risk of infectivity),^27^ the sensitivity of Ag-RDTs increased to 76·7% (95% CI 57·7 – 90·7) for the Abbott assay; 86·7% (69·3–96·3) for the Siemens Assay; 83·3% (65·3 – 94·4) for the Roche assay; 83·3% (65·3–94·4) for the Lepu assay; and to 70% (50·6–85·3%) for the Surescreen assay (Figure 2).

**Table 1.**
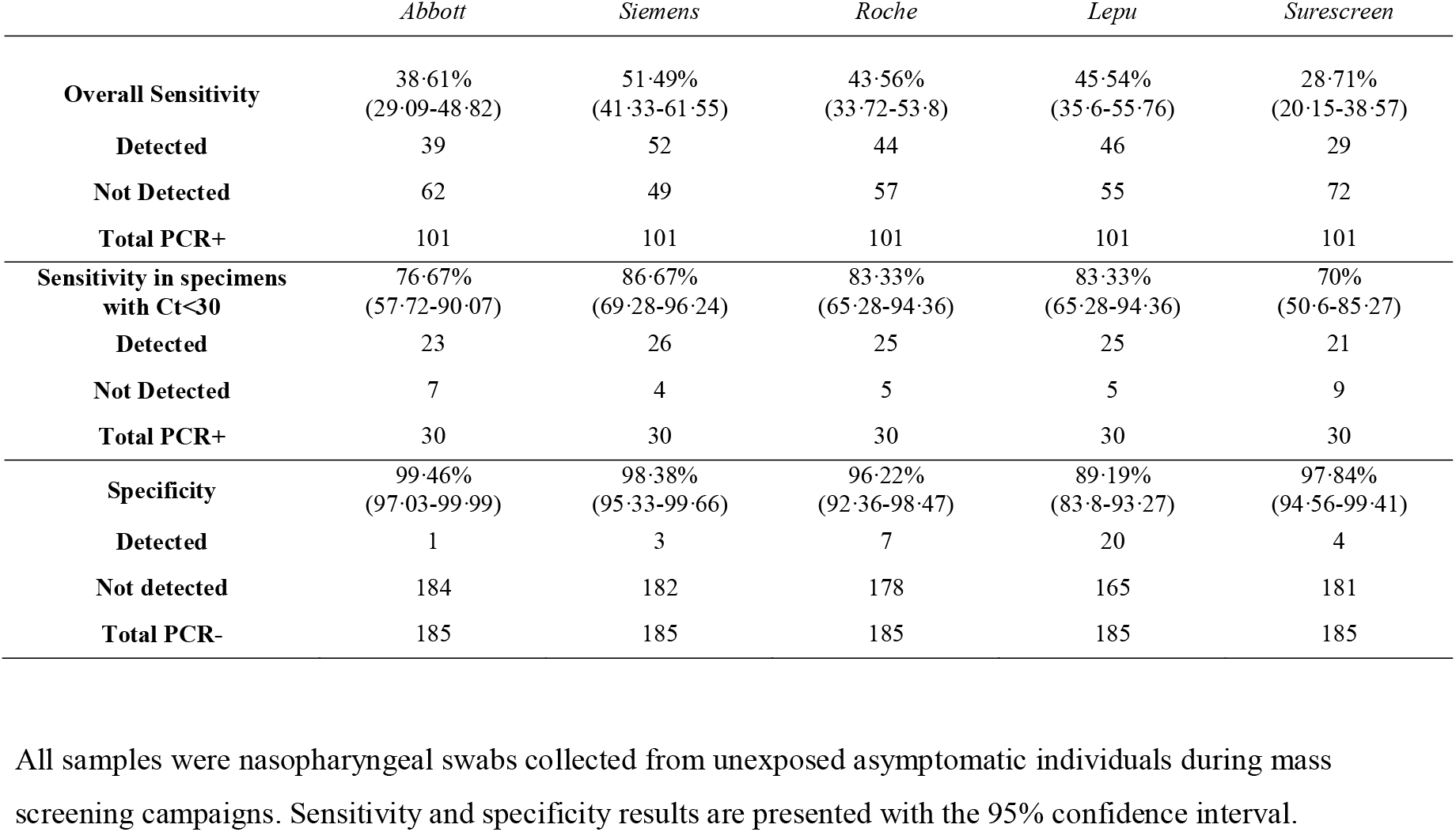
Sensitivity and specificity of the antigen-detecting rapid diagnostic tests for SARS-CoV-2.

**Figure 2.**
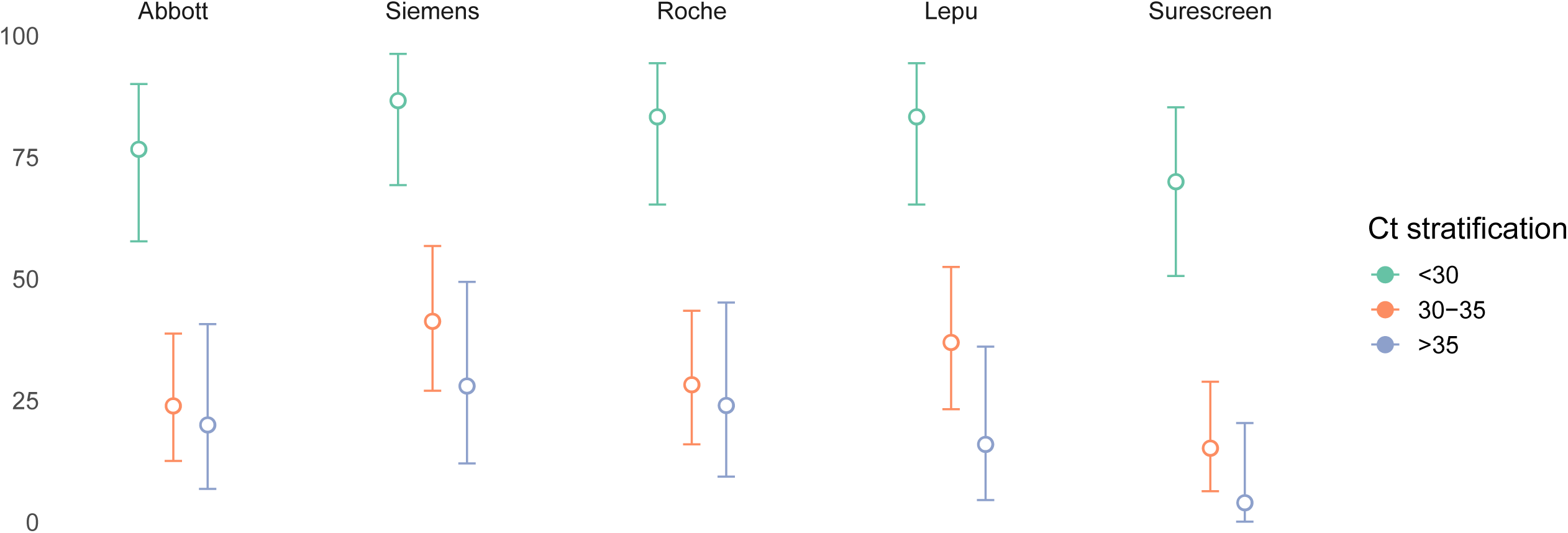
Sensitivity of the antigen-detecting rapid diagnostic tests according to the cycle threshold value of the RT-qPCR analysis. Bars show the 95% confidence interval of the estimated sensitivity.

Of the 286 samples analysed by Ag-RDTs, 222 (77·6%) had concordant results across all Ag-RDT assessed. The 29 samples with concordant positive results across Ag-RDTs were all PCR-positive. Conversely, 37 (19·2%) of 193 specimens with negative results in all Ag-RDTs were PCR positive. Figure 3 shows the distribution of Ag-RDT results in samples with discordant results. The Ag-RDT that most often yielded a positive result in samples with negative results in all other Ag-RDTs was the Lepu assay (n=23; 35·9%), followed by the Siemens assay (n=10, 15·6%). Table S2 summarizes the cycle threshold distribution across discordances.

**Figure 3.**
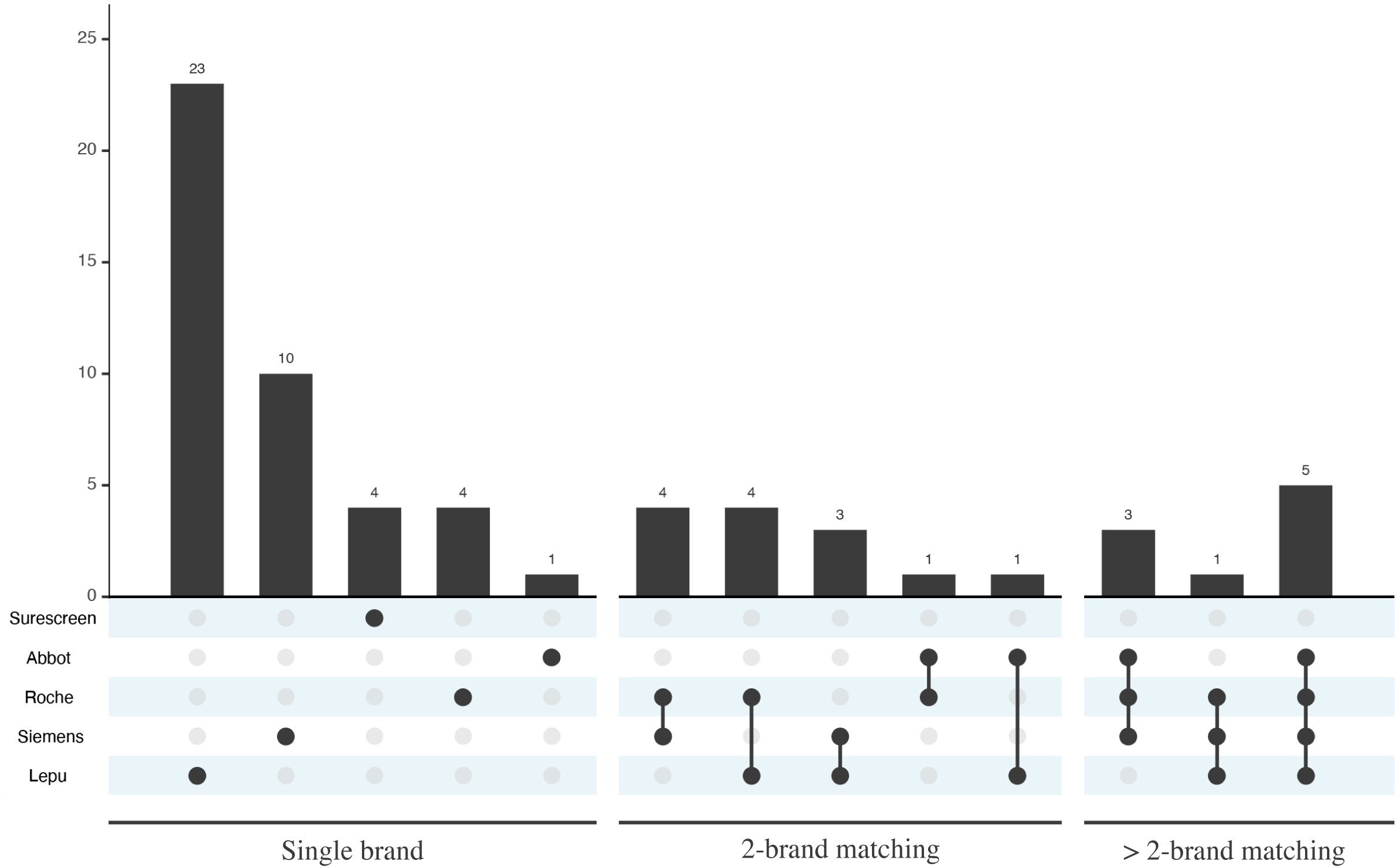
Discordance analysis between Ag-RDTs. Bars show the number of samples for each discordance pattern. Black dots and grey dots indicate the assays showing positive and negative results in each discordance pattern. Table S2 summarizes the cycle threshold distribution across discordances.

To provide an estimate of misidentified cases―either false-positive or false-negative cases―that can be used for making decisions in the public health setting, we modelled the positive and negative predictive value for a prevalence range consistent with a mass screening of unexposed asymptomatic individuals (Figure 4A). For the overall study sample, the estimated positive predictive value (PPV) at a 1% prevalence ranged from 4·1% to 41·9%, with the Lepu assay and the Abbott assay, respectively (Table S3). The estimated PPVs notably increased for the <30 Ct subgroup of samples (Figure 4B), and when prevalence in the population was higher. The estimated negative predictive value (NPV) at 1% prevalence ranged from 99·3% to 99·5%, with the Surescreen assay and the Siemens assay, respectively.

**Figure 4.**
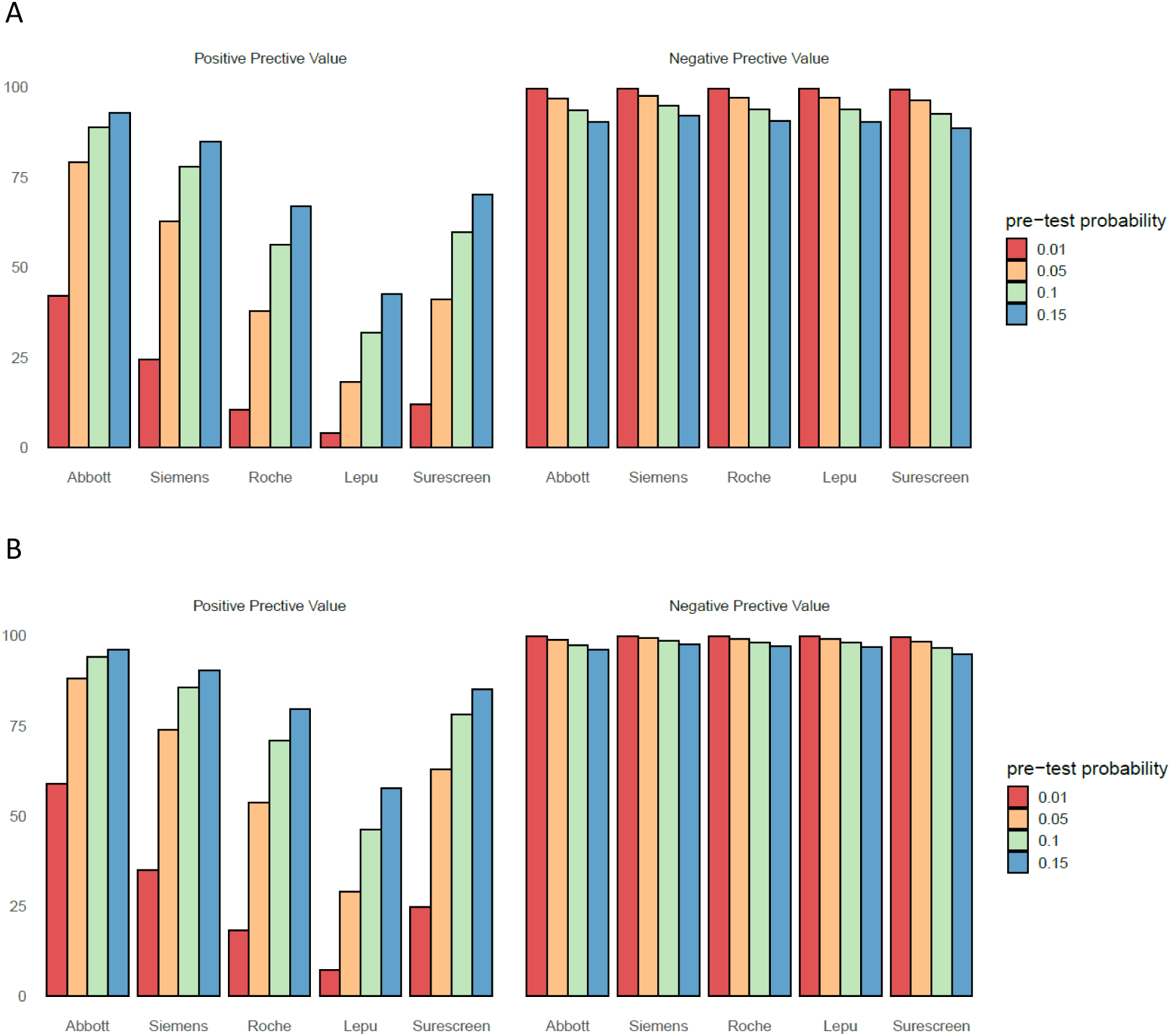
Positive Predictive Value and Negative Predictive Value according to pre-test probabilities. **A**: overall sample (n= 286). **B**: samples with cycle threshold <30 in the RT-qPCR assay. Table S3 provides detailed values and confidence intervals for predicted false negative and false positives in the investigated prevalence.

## Discussion

In this study, we compared head-to-head the sensitivity and specificity of five Ag-RDTs to screen SARS-CoV-2 infected individuals with unknown exposure and no clinical suspicion of COVID-19. Four of the tested Ag-RDTs (i.e., Abbott, Siemens, Roche, and Surescreen assays) showed a specificity higher than 96%. Regarding sensitivity, despite it was low for the overall sample (range 29% to 51%), the corresponding values for the subset of samples with a RT-qPCR value Ct <30 were higher than 80% for the Siemens, Roche, and Lepu assays. This finding is of particular interest for the proposed use of Ag-RDT as a reliable alternative to RT-qPCR for the rapid detection of individuals with higher risk of infectivity in mass screening of asymptomatic individuals. Pre-clinical studies have persistently reported a very low infectious capacity of respiratory specimens with viral loads below 10^6^ genome copies/mL, which usually correspond to a Ct of approximately 29 – 31.^4,7,28^ These findings align with the significant increase of the secondary attack rate for values of Ct <30,^29^ indicating higher infectiousness among individuals with viral loads below this Ct threshold.

Although sensitivity and specificity are important intrinsic characteristics of a test, the number of expected errors when using the test for screening purposes strongly depends on the prevalence of the infection in the screened sample. Hence, positive and negative predictive values are a mainstay for making public health decisions regarding the use of a test. The reported prevalence of SARS-CoV-2 infection in PCR-based untargeted screenings of the general population typically ranges between 1% and 3%, depending on the virus transmission context.^22,30^ In low prevalence settings, Ag-RDTs will have a high NPV but a low PPV. According to our estimate, the NPV for SARS-CoV-2 infections at 1% prevalence was higher than 99% for all test, suggesting that a negative test may not require confirmation. In contrast, the PPV at 1% prevalence was lower than 50% in all tests, suggesting that a positive result will need immediate confirmation by RT-qPCR, even for highly specific assays.

Our study has several strengths and limitations. We used the same fresh set of samples for assessing five different Ag-RDTs and the sample size met the FIND recommendation for retrospective assessments of the clinical performance of these tests. Furthermore, to our knowledge, this is the first head-to-head comparison of Ag-RDT in asymptomatic screenings, an intended use proposed by various authors.^4,9,16,22^ On the other hand, our study was limited by the small number of specimens with Ct <30, a threshold deemed of interest for the use of Ag-RDT in screenings of the general population. In our sample, specimens below this threshold accounted for 30%; however, other authors have reported proportions of nearly 60% in random screenings of the general population.^22^ Of note, we used specimens in transport medium. This approach is convenient for mass screening strategies in which individuals with positive Ag-RDT results may need further diagnostic confirmation by PCR. However, only one manufacturer (i.e., the Roche assay) provided instructions on how to process samples collected in virus transport medium. The consistency of our results across assays, particularly regarding negative results, suggests that the use of this media had a little or negligible impact on test performance. Finally, it is worth mentioning that all nasopharyngeal swabs in our analysis were collected by trained healthcare professionals. According to a recent report of lateral flow viral antigen detection devices, the positivity rate might be lower in screenings performed by non-trained people.^8^

Our results provide policymakers with evidence on the use of Ag-RDT for mass screening of unexposed, asymptomatic individuals. Two commercial, widely available assays can be used for SARS-CoV-2 antigen testing to achieve sensitivity in specimens with a Ct<30 and specificity of at least 80% and 96%, respectively. While these tests may overlook SARS-CoV-2 infection with low viral loads, they accurately detect individuals with high viral loads and, therefore, at higher risk of transmission. Our findings also support the idea that Ag-RDTs can be used for mass screening in low prevalence settings and accurately rule out a highly infectious case in such setting. In models according to population prevalence, all Ag-RDTs will have a NPV >99% and a PPV<50% at 1% prevalence. Together with the ease of use, low cost, and short turnaround time, this feature makes them an excellent tool for frequent mass screenings of asymptomatic people. In low-income countries with limited laboratory resources, the trade-off between targeted PCR analyses and massive screenings with Ag-RDTs should be carefully considered.

## Data Availability

Data are available upon request tp the corresponding author

## Contributors

OM, BB, IB designed the study. PR, BB, AEB, SS, ESA, AA, MU, MCM, PMM performed the laboratory procedures, and organized the data. BB, PR and DO verified the underlying data. DO did statistical analysis. BB, OM wrote the first draft with revisions and input from IB, QB, CGB, MVM, NP, JA, BC, MM. Funding acquisition by OM, IB, QB, JA, BC. All authors approved the final version.

## Declaration of interests

We declare no conflicts of interest.

## Acknowledgements

The authors would like to thank Gerard Carot-Sans (PhD) for providing professional medical writing support during the preparation of the manuscript. We thank Laia Comí, Josefa Gómez, Maria Pilar Rodríguez and Aida Sanz for technical support with samples selection and storing.

Bárbara Baro is a Beatriu de Pinós postdoctoral fellow granted by the Government of Catalonia’s Secretariat for Universities and Research, and by Marie Sklodowska-Curie Actions COFUND Programme (BP3, 801370)

ISGlobal receives support from the Spanish Ministry of Science and Innovation through the “Centro de Excelencia Severo Ochoa 2019-2023” Program (CEX2018-000806-S), and support from the Generalitat de Catalunya through the CERCA Program.

## Notes

### Competing Interest Statement

The authors have declared no competing interest.

### Funding Statement

Blueberry diagnostics, Fundacio Institut d'Investigacio en Ciencies de la Salut Germans Trias i Pujol, and #YoMeCorono.org crowdfunding campaign.

### Author Declarations

The study received approval from the Ethics Committe of the Hospital Universitari Germans Trias i Pujol

## References

1 Pavelka M, Van-Zandvoort K, Abbott S, et al. The effectiveness of population-wide, rapid antigen test based screening in reducing SARS-CoV-2 infection prevalence in Slovakia. medRxiv 2020. DOI:10.1101/2020.12.02.20240648.

2 World Health Organization (WHO). Diagnostic testing for SARS-CoV-2. https://www.who.int/publications/i/item/diagnostic-testing-for-sars-cov-2 (accessed Oct 6, 2020).

3 Gorzalski AJ, Tian H, Laverdure C, et al. High-Throughput Transcription-mediated amplification on the Hologic Panther is a highly sensitive method of detection for SARS-CoV-2. J Clin Virol 2020; 129: 104501.

4 Mina MJ, Parker R, Larremore DB. Rethinking COVID-19 test sensitivity—A strategy for containment. N Engl J Med 2020; 383: 10.1056/NEJMp2025631.

5 Wölfel R, Corman VM, Guggemos W, et al. Virological assessment of hospitalized patients with COVID-2019. Nature 2020; 581: 465–9.

6 La Scola B, Le Bideau M, Andreani J, et al. Viral RNA load as determined by cell culture as a management tool for discharge of SARS-CoV-2 patients from infectious disease wards. Eur J Clin Microbiol Infect Dis 2020; 39: 1059–61.

7 Quicke K, Gallichote E, Sexton N, et al. Longitudinal Surveillance for SARS-CoV-2 RNA Among Asymptomatic Staff in Five Colorado Skilled Nursing Facilities: Epidemiologic, Virologic and Sequence Analysis. medRxiv Prepr Serv Heal Sci 2020; : 2020.06.08.20125989.

8 PHE Porton Down & University of Oxford. Preliminary report from the Joint PHE Porton Down & University of Oxford SARS-CoV-2 test development and validation cell: Rapid evaluation of Lateral Flow Viral Antigen detection devices (LFDs) for mass community testing. https://www.ox.ac.uk/sites/files/oxford/media_wysiwyg/UKevaluation_PHEPortonDownUniversityofOxford_final.pdf (accessed Jan 3, 2020).

9 Larremore DB, Wilder B, Lester E, et al. Test sensitivity is secondary to frequency and turnaround time for COVID-19 screening. Sci Adv 2020; : eabd5393.

10 Porte L, Legarraga P, Vollrath V, et al. Evaluation of novel antigen-based rapid detection test for the diagnosis of SARS-CoV-2 in respiratory samples. Int J Infect Dis 2020; 99: 328–333.

11 Lambert-Niclot S, Cuffel A, Le Pape S, et al. Evaluation of a rapid diagnostic assay for detection of sars-cov-2 antigen in nasopharyngeal swabs. J Clin Microbiol 2020; 58: 0– 3.

12 Lindner AK, Nikolai O, Kausch F, et al. Head-to-head comparison of SARS-CoV-2 antigen-detecting rapid test with self-collected anterior nasal swab versus professional-collected nasopharyngeal swab. medRxiv. 2020; : 2003961.

13 Albert E, Torres I, Bueno F, et al. Field evaluation of a rapid antigen test (Panbio™ COVID-19 Ag Rapid Test Device) for COVID-19 diagnosis in primary healthcare centres. Clin Microbiol Infect 2020; 0. DOI:10.1016/j.cmi.2020.11.004.

14 Linares M, Pérez-Tanoira R, Carrero A, et al. Panbio antigen rapid test is reliable to diagnose SARS-CoV-2 infection in the first 7 days after the onset of symptoms. J Clin Virol 2020; 133: 104659.

15 Cerutti F, Burdino E, Milia MG, et al. Urgent need of rapid tests for SARS CoV-2 antigen detection: Evaluation of the SD-Biosensor antigen test for SARS-CoV-2. J Clin Virol 2020; 132: 104654.

16 Alemany A, Baró B, Ouchi D, et al. Analytical and clinical performance of the panbio COVID-19 antigen-detecting rapid diagnostic test. J Infect 2021; 18: 16.

17 Torres I, Poujois S, Albert E, Colomina J, Navarro D. Evaluation of a rapid antigen test (Panbio™COVID-19 Ag rapid test device) for SARS-CoV-2 detection in asymptomatic close contacts of COVID-19 patients. Clin Microbiol Infect 2021; •: 1.

18 Weitzel T, Legarraga P, Iruretagoyena M, et al. Comparative evaluation of four rapid SARS-CoV-2 antigen detection tests using universal transport medium. Travel Med Infect Dis 2021; 39: 101942.

19 Corman VM, Haage VC, Bleicker T, et al. Comparison of seven commercial SARS-CoV-2 rapid Point-of-Care Antigen tests. medRxiv. 2020; : 2020.11.12.20230292.

20 Foundation for Innovative New Diagnostics. Comparative evaluation of lateral flow assay tests that directly detect antigens of SARS-CoV-2. https://www.finddx.org/wp-content/uploads/2020/04/20200421-COVID-Ag-RDT-Evaluation-Synopsis.pdf (accessed Dec 20, 2020).

21 Seegene Inc. Allplex™ 2019-nCoV Assay. https://www.seegene.com/assays/allplex_2019_ncov_assay (accessed Dec 20, 2020).

22 Pilarowski G, Lebel P, Sunshine S, et al. Performance characteristics of a rapid SARS-CoV-2 antigen detection assay at a public plaza testing site in San Francisco. medRxiv 2020; published online Nov 12. DOI:10.1101/2020.11.02.20223891.

23 Altman DG, Bland JM. Statistics Notes: Diagnostic tests 1: Sensitivity and specificity. BMJ 1994; 308: 1552.

24 Altman DG, Bland j. M. Statistics Notes: Diagnostic tests 2: Predictive values. BMJ 1994; 309: 102.

25 Collett D. Modelling Binary Data, Second. Chapman and Hall/CRC, 2002 https://www.routledge.com/Modelling-Binary-Data/Collett/p/book/9781584883241 (accessed Oct 22, 2020).

26 R Core Team. R: A language and environment for statistical com_puting. R Found. Stat. Comput. Vienna, Austria. 2017. https://www.r-project.org (accessed May 25, 2020).

27 World Health Organization (WHO). Antigen-detection in the diagnosis of SARS-CoV-2 infection using rapid immunoassays. 2020. https://www.who.int/publications/i/item/antigen-detection-in-the-diagnosis-of-sars-cov-2infection-using-rapid-immunoassays (accessed Sept 29, 2020).

28 Singanayagam A, Patel M, Charlett A, et al. Duration of infectiousness and correlation with RT-PCR cycle threshold values in cases of COVID-19, England, January to May 2020. Eurosurveillance 2020; 25: 2001483.

29 Marks M, Millat P, Ouchi D, et al. Transmission of Covid-19 in 282 clusters in Catalonia, Spain: a cohort study. Lancet Infect Dis 2020; : Manuscript accepted (in Press).

30 Oran DP, Topol EJ. Prevalence of Asymptomatic SARS-CoV-2 Infection_: A Narrative Review. Ann Intern Med 2020; 173: 362–7.

