## Supplementary materials for "Performance characteristics of five antigen-detecting rapid diagnostic test (Ag-RDT) for SARS-CoV-2 asymptomatic infection: a head-to-head benchmark comparison"

**Table S1. Main characteristics of five rapid SARS-CoV-2 rapid antigen tests (according to the manufacturer)**

| **Assay name** | PanBio^TM^ COVID-19 Ag Rapid Test Device | CLINITEST®  Rapid COVID-19 Antigen Test | SARS-CoV-2 Rapid Antigen Test | SARS-CoV-2 Antigen Rapid Test Kit | COVID-19 Antigen Test Cassette |
| --- | --- | --- | --- | --- | --- |
| **Distributor** | Abbott Rapid  Diagnostics | Siemens Healthineers (Shangai International Holding Corp) | Roche Diagnostics | Lepu Medical (Europe) cooperatief U.A. | SureScreen Diagnostics |
| **Manufacturer** | Panbio Ltd. | Healgen Scientific Limited Liability Company | SD Biosensor | Beijing Lepu Medical Technology Co., Ltd. | SureScreen Diagnostics Ltd |
| **Country of origin** | USA | USA | Republic of Korea | P.R. China | UK |
| **Certification** | CE-IVD | CE-IVD | CE-IVD | CE-IVD | CE-IVD |
| **Specimen** | Nasopharyngeal swab | Nasopharyngeal or nasal swab | Nasopharyngeal swab | Nasopharyngeal or oropharyngeal swab | Nasopharyngeal or oropharyngeal swab |
| **Intended use** | Suspected cases of COVID-19 (clinical and/or epidemiological criteria) | Suspected cases of COVID-19 | Suspected cases of COVID-19 | Clinical samples | Suspected cases of COVID-19 within first 2weeks of symptoms |
| **Limit of detection**  **(manufacturer)** | 2,5 X10^1,8^ TCID50/mL | 1,2 x 10^2^ TCID50/mL | 3,1 X 10^2,2^ TCID50/mL | Not specified | 2x10 ^2,4^  TCID50/mL |
| **Reported sensitivity and specificity** | 97.9% (95.2-99.3%)  99.4% (97.0-100.0%) | 96.7% (88.7-99.60%)  99.22% (97.21-99.91%) | 96.52% (91.33-99.04%)  99.68% (98.22-99.99%) | No less than 90%  No less than 100% | 94.55% (83.93-98.58%)  100% (97.1-100%) |
| **Format** | Cassette | Cassette | Cassette | Card | Cassette |
| **Volume in extraction tube** | Marked line (~300uL) | 10 drops | Aliquoted (~250uL) | NA | 10 drops |
| **Volume applied**  **into cassette** | 5 drops | 4 drops | 3 drops | 6 drops | 2 drops |
| **Incubation** | 15-20 minutes | 15-20 minutes | 15-30 minutes | 15-20 minutes | 15 minutes |
| **Readout** | Visual: colored band | Visual: colored band | Visual: colored band | Visual: colored band | Visual: colored band |
| **UTM protocol** | Not specified | Not specified | Reported | Not specified | Not specified |
| **Cross-reactivity against other human virus** | Reported | Reported | Reported | Reported | Reported |

**Table S2.** Discordance analysis

|  |  |  |  |  |  |  |  | **Cases** | | |
| --- | --- | --- | --- | --- | --- | --- | --- | --- | --- | --- |
| **Abbott** | **Siemens** | **Roche** | **Lepu** | **SureScreen** | **All samples** | **PCR+** | **%PCR+** | **<30** | **30-35** | **>35** |
| **Full agreement** | | | | | | | | | | |
| + | + | + | + | + | 29 | 29 | 100 | 21 | 7 | 1 |
| - | - | - | - | - | 193 | 37 | 19.17 | 3 | 20 | 14 |
| **Discordance** | | | | | | | | | | |
| - | - | - | + | - | 23 | 8 | 34.78 | 1 | 6 | 1 |
| - | + | - | - | - | 10 | 8 | 80 |  | 5 | 3 |
| - | - | + | - | - | 4 | 2 | 50 |  |  | 2 |
| + | - | - | - | - | 1 | 1 | 100 |  |  | 1 |
| - | - | - | - | + | 4 | 0 | 0 |  |  |  |
| - | + | - | + | - | 3 | 3 | 100 | 1 | 2 |  |
| - | + | + | - | - | 4 | 3 | 75 | 1 | 2 |  |
| - | - | + | + | - | 4 | 0 | 0 |  |  |  |
| + | - | - | + | - | 1 | 0 | 0 |  |  |  |
| - | + | + | + | - | 1 | 1 | 100 | 1 |  |  |
| + | + | + | - | - | 3 | 3 | 100 | 1 | 1 | 1 |
| + | + | + | + | - | 5 | 5 | 100 | 1 | 2 | 2 |
| + | - | + | - | - | 1 | 1 | 100 |  | 1 |  |

**Table S3.** Estimated false negative and false positives from the positive- and negative-predictive values modeled for each prevalence.

|  |  |  | | |  | | |  | | |  | | |  | **All samples** | | | **Samples CT<30** | | | **Samples CT [30-35]** | | | **Samples CT>35** | | |
| --- | --- | --- | --- | --- | --- | --- | --- | --- | --- | --- | --- | --- | --- | --- | --- | --- | --- | --- | --- | --- | --- | --- | --- | --- | --- | --- |
|  | **Prevalence** | **Specificity** | | **Sensitivity** | | | **FN** | | | **FP** | | **Total**  **Errors** | | | **PPV** | **NPV** | | **PPV** | **NPV** | | **PPV** | **NPV** | | **PPV** | **NPV** | |
| Abbott | 0.01 | 99.46%  (97.03-99.99) | 38.61%  (29.09-48.82) | | | 6  (5-7) | | | 5  (0-30) | | | | 12  (5-37) | | 41.91  (8.99-97.3) | | 99.38  (99.27-99.49) | 58.89  (16.39-98.52) | | 99.76  (99.56-99.9) | 30.88  (4.1-96.62) | | 99.23  (99.1-99.39) | 27.21  (2.27-96.78) | | 99.19  (99.04-99.4) |
| Siemens | 0.01 | 98.38%  (95.33-99.66) | 51.49%  (41.33-61.55) | | | 5  (4-6) | | | 16  (3-47) | | | | 21  (7-53) | | 24.28  (8.21-64.94) | | 99.5  (99.38-99.61) | 35.06  (13.04-74.33) | | 99.86  (99.68-99.96) | 20.46  (5.52-63.08) | | 99.4  (99.23-99.56) | 14.85  (2.55-59.78) | | 99.27  (99.08-99.49) |
| Roche | 0.01 | 96.22%  (92.36-98.47) | 43.56%  (33.72-53.8) | | | 6  (5-7) | | | 38  (16-77) | | | | 44  (20-84) | | 10.42  (4.27-26.15) | | 99.41  (99.28-99.53) | 18.2  (7.94-38.31) | | 99.83  (99.62-99.94) | 7.02  (2.07-22.24) | | 99.25  (99.09-99.42) | 6.02  (1.22-22.9) | | 99.21  (99.02-99.44) |
| Lepu | 0.01 | 89.19%  (83.8-93.27) | 45.54%  (35.6-55.76) | | | 5  (4-6) | | | 109  (68-164) | | | | 115  (72-170) | | 4.08  (2.17-7.72) | | 99.39  (99.23-99.52) | 7.22  (3.91-12.41) | | 99.81  (99.58-99.94) | 3.34  (1.43-7.3) | | 99.29  (99.08-99.49) | 1.47  (0.28-5.14) | | 99.06  (98.86-99.31) |
| Surescreen | 0.01 | 97.84%  (94.56-99.41) | 28.71%  (20.15-38.57) | | | 7  (6-8) | | | 22  (6-55) | | | | 29  (12-63) | | 11.83  (3.6-39.68) | | 99.27  (99.15-99.38) | 24.64  (8.58-59.26) | | 99.69  (99.48-99.85) | 6.64  (1.16-32.99) | | 99.13  (99.01-99.28) | 1.83  (0.02-25.77) | | 99.02  (98.94-99.2) |
| Abbott | 0.05 | 99.46%  (97.03-99.99) | 38.61%  (29.09-48.82) | | | 31  (26-35) | | | 6  (0-31) | | | | 36  (26-67) | | 78.99  (33.98-99.47) | | 96.85  (96.3-97.38) | 88.19  (50.52-99.71) | | 98.78  (97.76-99.48) | 69.96  (18.21-99.33) | | 96.13  (95.47-96.88) | 66.07  (10.78-99.37) | | 95.94  (95.19-96.97) |
| Siemens | 0.05 | 98.38%  (95.33-99.66) | 51.49%  (41.33-61.55) | | | 24  (19-29) | | | 17  (4-49) | | | | 41  (23-78) | | 62.56  (31.8-90.61) | | 97.47  (96.86-98.01) | 73.77  (43.87-93.79) | | 99.29  (98.33-99.8) | 57.28  (23.34-89.9) | | 96.96  (96.13-97.77) | 47.61  (11.99-88.56) | | 96.29  (95.37-97.4) |
| Roche | 0.05 | 96.22%  (92.36-98.47) | 43.56%  (33.72-53.8) | | | 28  (23-33) | | | 40  (16-80) | | | | 68  (39-114) | | 37.73  (18.85-64.85) | | 97.01  (96.36-97.59) | 53.69  (31.02-76.39) | | 99.1  (98.06-99.7) | 28.22  (9.92-59.85) | | 96.22  (95.43-97.07) | 25.03  (6.06-60.75) | | 96.01  (95.09-97.15) |
| Lepu | 0.05 | 89.19%  (83.8-93.27) | 45.54%  (35.6-55.76) | | | 27  (22-32) | | | 114  (71-171) | | | | 141  (93-203) | | 18.15  (10.37-30.36) | | 96.89  (96.11-97.56) | 28.86  (17.5-42.46) | | 99.03  (97.87-99.68) | 15.25  (7.01-29.09) | | 96.41  (95.4-97.39) | 7.23  (1.45-22.01) | | 95.28  (94.34-96.52) |
| Surescreen | 0.05 | 97.84%  (94.56-99.41) | 28.71%  (20.15-38.57) | | | 36  (31-40) | | | 23  (6-57) | | | | 58  (37-97) | | 41.14  (16.3-77.42) | | 96.31  (95.74-96.85) | 63.02  (32.85-88.34) | | 98.41  (97.32-99.23) | 27.03  (5.78-71.96) | | 95.64  (95.05-96.37) | 8.87  (0.1-64.4) | | 95.09  (94.73-95.95) |
| Abbott | 0.1 | 99.46%  (97.03-99.99) | 38.61%  (29.09-48.82) | | | 61  (51-71) | | | 6  (0-33) | | | | 67  (51-104) | | 88.81  (52.07-99.75) | | 93.58  (92.49-94.62) | 94.03  (68.31-99.86) | | 97.46  (95.38-98.91) | 83.1  (31.98-99.68) | | 92.17  (90.9-93.63) | 80.43  (20.33-99.7) | | 91.8  (90.36-93.82) |
| Siemens | 0.1 | 98.38%  (95.33-99.66) | 51.49%  (41.33-61.55) | | | 49  (38-59) | | | 18  (4-52) | | | | 67  (42-111) | | 77.91  (49.6-95.32) | | 94.81  (93.6-95.89) | 85.59  (62.26-96.96) | | 98.52  (96.54-99.58) | 73.89  (39.13-94.95) | | 93.78  (92.16-95.4) | 65.74  (22.33-94.24) | | 92.48  (90.7-94.66) |
| Roche | 0.1 | 96.22%  (92.36-98.47) | 43.56%  (33.72-53.8) | | | 56  (46-66) | | | 42  (17-85) | | | | 98  (63-151) | | 56.13  (32.9-79.57) | | 93.88  (92.62-95.04) | 70.99  (48.7-87.23) | | 98.11  (95.99-99.37) | 45.35  (18.86-75.88) | | 92.35  (90.82-94) | 41.34  (11.98-76.57) | | 91.93  (90.17-94.17) |
| Lepu | 0.1 | 89.19%  (83.8-93.27) | 45.54%  (35.6-55.76) | | | 54  (44-64) | | | 120  (75-180) | | | | 175  (119-244) | | 31.88  (19.62-47.93) | | 93.65  (92.13-94.99) | 46.13  (30.92-60.91) | | 97.97  (95.6-99.33) | 27.53  (13.73-46.41) | | 92.72  (90.76-94.64) | 14.12  (3.02-37.33) | | 90.53  (88.76-92.92) |
| Surescreen | 0.1 | 97.84%  (94.56-99.41) | 28.71%  (20.15-38.57) | | | 71  (61-80) | | | 24  (7-60) | | | | 95  (68-140) | | 59.6  (29.14-87.86) | | 92.51  (91.42-93.57) | 78.25  (50.81-94.12) | | 96.71  (94.51-98.38) | 43.88  (11.47-84.42) | | 91.22  (90.09-92.64) | 17.05  (0.21-79.25) | | 90.17  (89.49-91.83) |
| Abbott | 0.15 | 99.46%  (97.03-99.99) | 38.61%  (29.09-48.82) | | | 92  (77-106) | | | 6  (0-35) | | | | 98  (77-141) | | 92.65  (63.31-99.84) | | 90.18  (88.58-91.72) | 96.16  (77.4-99.91) | | 96.02  (92.86-98.28) | 88.65  (42.75-99.8) | | 88.11  (86.28-90.25) | 86.72  (28.84-99.81) | | 87.57  (85.51-90.53) |
| Siemens | 0.15 | 98.38%  (95.33-99.66) | 51.49%  (41.33-61.55) | | | 73  (58-88) | | | 19  (4-55) | | | | 92  (62-143) | | 84.85  (60.99-97) | | 91.99  (90.2-93.63) | 90.41  (72.38-98.06) | | 97.66  (94.62-99.34) | 81.8  (50.52-96.76) | | 90.47  (88.1-92.89) | 75.29  (31.35-96.29) | | 88.56  (86-91.78) |
| Roche | 0.15 | 96.22%  (92.36-98.47) | 43.56%  (33.72-53.8) | | | 85  (69-99) | | | 45  (18-90) | | | | 129  (87-189) | | 67.02  (43.78-86.08) | | 90.62  (88.76-92.35) | 79.54  (60.12-91.56) | | 97.03  (93.78-99) | 56.86  (26.97-83.33) | | 88.37  (86.17-90.8) | 52.82  (17.77-83.84) | | 87.77  (85.24-91.05) |
| Lepu | 0.15 | 89.19%  (83.8-93.27) | 45.54%  (35.6-55.76) | | | 82  (66-97) | | | 127  (79-191) | | | | 209  (146-287) | | 42.64  (27.94-59.38) | | 90.27  (88.06-92.28) | 57.63  (41.56-71.22) | | 96.81  (93.19-98.94) | 37.63  (20.18-57.9) | | 88.91  (86.08-91.75) | 20.71  (4.71-48.62) | | 85.75  (83.26-89.21) |
| Surescreen | 0.15 | 97.84%  (94.56-99.41) | 28.71%  (20.15-38.57) | | | 107  (92-120) | | | 25  (7-64) | | | | 132  (99-184) | | 70.09  (39.51-92) | | 88.61  (87.03-90.17) | 85.1  (62.13-96.21) | | 94.87  (91.56-97.45) | 55.4  (17.06-89.59) | | 86.74  (85.12-88.79) | 24.61  (0.33-85.85) | | 85.24  (84.29-87.61) |
| Abbott | 0.2 | 99.46%  (97.03-99.99) | 38.61%  (29.09-48.82) | | | 123  (102-142) | | | 7  (0-37) | | | | 130  (103-179) | | 94.7  (70.97-99.89) | | 86.63  (84.55-88.66) | 97.26  (82.91-99.94) | | 94.46  (90.18-97.58) | 91.71  (51.4-99.86) | | 83.95  (81.62-86.72) | 90.24  (36.47-99.87) | | 83.26  (80.64-87.09) |
| Siemens | 0.2 | 98.38%  (95.33-99.66) | 51.49%  (41.33-61.55) | | | 97  (77-117) | | | 20  (4-58) | | | | 117  (81-176) | | 88.81  (68.89-97.87) | | 89.02  (86.67-91.2) | 93.04  (78.78-98.62) | | 96.72  (92.54-99.07) | 86.43  (59.13-97.69) | | 87.02  (83.93-90.22) | 81.19  (39.28-97.35) | | 84.53  (81.26-88.73) |
| Roche | 0.2 | 96.22% (92.36-98.47) | 43.56%  (33.72-53.8) | | | 113  (92-133) | | | 47  (19-96) | | | | 160  (112-228) | | 74.22  (52.46-89.76) | | 87.21  (84.79-89.5) | 84.63  (68.11-93.89) | | 95.85  (91.41-98.59) | 65.12  (34.34-87.62) | | 84.29  (81.47-87.45) | 61.33  (23.44-88.03) | | 83.51  (80.3-87.77) |
| Lepu | 0.2 | 89.19%  (83.8-93.27) | 45.54%  (35.6-55.76) | | | 109  (88-129) | | | 135  (84-203) | | | | 244  (173-331) | | 51.3  (35.46-67.44) | | 86.76  (83.88-89.4) | 65.84  (50.18-77.8) | | 95.54  (90.61-98.51) | 46.08  (26.37-66.08) | | 84.98  (81.36-88.7) | 27.01  (6.54-57.27) | | 80.94  (77.83-85.37) |
| Surescreen | 0.2 | 97.84%  (94.56-99.41) | 28.71%  (20.15-38.57) | | | 143  (123-160) | | | 27  (7-68) | | | | 170  (130-228) | | 76.85  (48.06-94.21) | | 84.59  (82.57-86.62) | 89  (69.92-97.3) | | 92.88  (88.45-96.43) | 63.76  (22.56-92.42) | | 82.19  (80.15-84.83) | 31.62  (0.46-89.57) | | 80.3  (79.11-83.31) |
| Abbott | 0.5 | 99.46%  (97.03-99.99) | 38.61%  (29.09-48.82) | | | 307  (256-355) | | | 11  (0-59) | | | | 318  (256-414) | | 98.62  (90.72-99.97) | | 61.84  (57.78-66.14) | 99.3  (95.1-99.98) | | 81  (69.65-90.96) | 97.79  (80.88-99.96) | | 56.66  (52.61-62.02) | 97.37  (69.66-99.97) | | 55.42  (51.01-62.77) |
| Siemens | 0.5 | 98.38%  (95.33-99.66) | 51.49%  (41.33-61.55) | | | 243  (192-293) | | | 32  (7-93) | | | | 275  (199-387) | | 96.95  (89.86-99.46) | | 66.97  (61.9-72.16) | 98.16  (93.69-99.65) | | 88.06  (75.63-96.37) | 96.22  (85.26-99.41) | | 62.63  (56.63-69.75) | 94.53  (72.12-99.32) | | 57.74  (52.02-66.32) |
| Roche | 0.5 | 96.22%  (92.36-98.47) | 43.56%  (33.72-53.8) | | | 282  (231-331) | | | 76  (31-153) | | | | 358  (262-484) | | 92.01  (81.53-97.23) | | 63.03  (58.22-68.06) | 95.66  (89.52-98.4) | | 85.24  (72.68-94.58) | 88.19 (67.66-96.59) | | 57.29  (52.37-63.52) | 86.38  (55.05-96.71) | | 55.87  (50.47-64.22) |
| Lepu | 0.5 | 89.19%  (83.8-93.27) | 45.54%  (35.6-55.76) | | | 272  (221-322) | | | 216  (135-324) | | | | 488  (356-646) | | 80.82  (68.72-89.23) | | 62.09  (56.54-67.83) | 88.52  (80.12-93.34) | | 84.26  (70.7-94.3) | 77.37  (58.89-88.63) | | 58.59  (52.18-66.23) | 59.68  (21.88-84.28) | | 51.5  (46.75-59.34) |
| Surescreen | 0.5 | 97.84%  (94.56-99.41) | 28.71%  (20.15-38.57) | | | 356  (307-399) | | | 43  (12-109) | | | | 400  (319-508) | | 93  (78.73-98.49) | | 57.85  (54.21-61.81) | 97  (90.29-99.31) | | 76.53  (65.69-87.09) | 87.56  (53.82-97.99) | | 53.57  (50.24-58.29) | 64.91  (1.83-97.17) | | 50.47  (48.63-55.52) |
| Abbott | 0.85 | 99.46%  (97.03-99.99) | 38.61%  (29.09-48.82) | | | 522  (435-603) | | | 36  (1-198) | | | | 558  (436-801) | | 99.75  (98.23-100) | | 22.23  (19.45-25.64) | 99.88  (99.1-100) | | 42.93  (28.82-63.98) | 99.6  (96-99.99) | | 18.74  (16.38-22.37) | 99.53  (92.86-99.99) | | 17.99  (15.52-22.93) |
| Siemens | 0.85 | 98.38%  (95.33-99.66) | 51.49%  (41.33-61.55) | | | 412  (327-499) | | | 108  (22-311) | | | | 520  (349-810) | | 99.45  (98.05-99.9) | | 26.35  (22.28-31.39) | 99.67  (98.83-99.94) | | 56.56  (35.38-82.4) | 99.31  (97.04-99.9) | | 22.83  (18.73-28.92) | 98.99  (93.61-99.88) | | 19.43  (16.06-25.79) |
| Roche | 0.85 | 96.22%  (92.36-98.47) | 43.56%  (33.72-53.8) | | | 480  (393-563) | | | 252  (102-509) | | | | 732  (495-1073) | | 98.49  (96.16-99.5) | | 23.13  (19.74-27.33) | 99.21  (97.98-99.71) | | 50.46  (31.95-75.49) | 97.69  (92.22-99.38) | | 19.14  (16.25-23.51) | 97.29  (87.4-99.4) | | 18.26  (15.24-24.05) |
| Lepu | 0.85 | 89.19%  (83.8-93.27) | 45.54%  (35.6-55.76) | | | 463  (376-547) | | | 721  (449-1080) | | | | 1184  (825-1628) | | 95.98  (92.57-97.91) | | 22.42  (18.67-27.12) | 97.76  (95.8-98.76) | | 48.57  (29.87-74.47) | 95.09  (89.03-97.79) | | 19.98  (16.15-25.71) | 89.35  (61.35-96.81) | | 15.78  (13.41-20.48) |
| Surescreen | 0.85 | 97.84%  (94.56-99.41) | 28.71%  (20.15-38.57) | | | 606  (522-679) | | | 144  (39-363) | | | | 750  (562-1042) | | 98.69  (95.45-99.73) | | 19.5  (17.28-22.21) | 99.46  (98.14-99.88) | | 36.53  (25.25-54.35) | 97.55  (86.85-99.64) | | 16.92  (15.12-19.78) | 91.29  (9.53-99.49) | | 15.24  (14.31-18.05) |
| Abbott | 0.9 | 99.46%  (97.03-99.99) | 38.61%  (29.09-48.82) | | | 552  (461-638) | | | 54  (1-297) | | | | 607  (462-936) | | 99.84  (98.88-100) | | 15.26  (13.2-17.84) | 99.92  (99.43-100) | | 32.14  (20.32-52.79) | 99.75  (97.44-100) | | 12.68  (10.98-15.36) | 99.7  (95.38-100) | | 12.14  (10.37-15.78) |
| Siemens | 0.9 | 98.38%  (95.33-99.66) | 51.49%  (41.33-61.55) | | | 437  (346-528) | | | 162  (34-467) | | | | 599  (380-995) | | 99.65  (98.76-99.94) | | 18.39  (15.29-22.36) | 99.79  (99.26-99.96) | | 45.05  (25.64-74.68) | 99.57  (98.12-99.93) | | 15.7  (12.67-20.39) | 99.36  (95.88-99.92) | | 13.18  (10.75-17.95) |
| Roche | 0.9 | 96.22%  (92.36-98.47) | 43.56%  (33.72-53.8) | | | 508  (416-597) | | | 378  (153-764) | | | | 886  (569-1361) | | 99.04  (97.54-99.68) | | 15.93  (13.41-19.15) | 99.5  (98.72-99.82) | | 39.08  (22.81-65.98) | 98.53  (94.96-99.61) | | 12.97  (10.89-16.21) | 98.28  (91.68-99.62) | | 12.33  (10.17-16.62) |
| Lepu | 0.9 | 89.19%  (83.8-93.27) | 45.54%  (35.6-55.76) | | | 490  (398-580) | | | 1081  (673-1620) | | | | 1571  (1071-2200) | | 97.43  (95.19-98.68) | | 15.4  (12.63-18.98) | 98.58  (97.32-99.21) | | 37.29  (21.15-64.75) | 96.85  (92.8-98.59) | | 13.58  (10.81-17.9) | 93.02  (71.6-97.97) | | 10.55  (8.89-13.95) |
| Surescreen | 0.9 | 97.84%  (94.56-99.41) | 28.71%  (20.15-38.57) | | | 642  (553-719) | | | 216  (59-544) | | | | 858  (612-1263) | | 99.17  (97.09-99.83) | | 13.23  (11.63-15.24) | 99.66  (98.82-99.92) | | 26.6  (17.54-42.84) | 98.45  (91.3-99.77) | | 11.36  (10.09-13.44) | 94.33  (14.34-99.68) | | 10.17  (9.52-12.18) |
